# A functionally distinct neutrophil landscape in severe COVID-19 reveals opportunities for adjunctive therapies

**DOI:** 10.1101/2021.06.07.21258484

**Authors:** Rachita Panda, Fernanda Vargas E Silva Castanheira, Jared Schlechte, Bas GJ Surewaard, Hanjoo Brian Shim, Amanda Z Zucoloto, Zdenka Slavikova, Bryan G Yipp, Paul Kubes, Braedon McDonald

## Abstract

Acute respiratory distress syndrome (ARDS) is a life-threatening syndrome of respiratory failure and diffuse alveolar damage that results from dysregulated local and systemic immune activation, causing pulmonary vascular, parenchymal and alveolar damage. SARS-CoV-2 infection has become the dominant cause of ARDS worldwide, and emerging evidence implicates neutrophils and their cytotoxic arsenal of effector functions as central drivers of immune-mediated lung injury in COVID-19 ARDS. However, a key outstanding question is whether COVID-19 drives a unique program of neutrophil activation or effector functions that contributes to the severe pathogenesis of this pandemic illness, and whether this unique neutrophil response can be targeted to attenuate disease. Using a combination of high-dimensional single cell analysis and *ex vivo* functional assays of neutrophils from patients with COVID-19 ARDS compared to non-COVID ARDS (caused by bacterial pneumonia), we identified a functionally distinct landscape of neutrophil activation in COVID-19 ARDS that was intrinsically programmed during SARS-CoV-2 infection. Furthermore, neutrophils in COVID-19 ARDS were functionally primed to produce high amounts of neutrophil extracellular traps (NETs). Surprisingly, this unique pathological program of neutrophil priming escaped conventional therapy with dexamethasone, thereby revealing a promising target for adjunctive immunotherapy in severe COVID-19.

## INTRODUCTION

Acute respiratory distress syndrome (ARDS) is a life-threatening syndrome of hypoxemic respiratory failure and diffuse alveolar damage that results from an aberrant host response to respiratory and systemic insults, with the most common cause being severe pneumonia (1). Dysregulated local and systemic inflammation triggers widespread pulmonary vascular, parenchymal and alveolar damage, resulting in failure of gas exchange and critical illness (2). The mainstay of treatment for ARDS is supportive care including the use of mechanical ventilation, but ongoing research efforts strive to define the mechanisms of pathogenesis to enable targeted therapeutic interventions (2).

SARS-CoV-2 infection has emerged as the dominant cause of ARDS worldwide in the context of the COVID-19 pandemic (3). This has created an explosion of research to understand why SARS-CoV-2 infection is so prone to precipitating ARDS, and much has been learned about the pathogenesis including aberrant local and systemic immune responses elicited by the virus. Transcriptomic and proteomic analysis of the immune landscape in the bloodstream as well as lung tissue have revealed that COVID-19 is associated with a dysregulated myeloid cellular response, and in particular, that the progression towards severe disease is heralded by aberrant neutrophil activation (4–8). Autopsy studies of lung tissue from fatal COVID-19 have shown evidence of neutrophil infiltration within the pulmonary microvasculature and alveoli, albeit to a lesser extent than ARDS caused by bacterial pneumonia, suggesting distinct mechanisms of neutrophil responses to SARS-CoV-2 infection compared to other causes of ARDS (9, 10). Furthermore, neutrophils within the pulmonary microvasculature of severe COVID-19 are associated with widespread neutrophil extracellular trap (NETs) production (11–14). This dysregulated neutrophil effector response is thought to contribute to impaired gas exchange through the induction of microvascular thrombosis and perfusion defects, as well as tissue injury and diffuse alveolar damage.

Given the growing appreciation that immune-mediated pathology is the primary driver of disease in COVID-19 ARDS, it has become the focus of therapeutic intervention. Currently, the most established immunomodulatory therapy for severe COVID-19 is dexamethasone, which has been shown to reduce mortality in moderate to severe COVID-19 (15). However, the mechanisms by which dexamethasone improves clinical outcomes in COVID-19 are incompletely understood, and therefore, opportunities may exist for adjunctive immunomodulation directed at mechanisms that escape the activity of dexamethasone.

A key outstanding question is whether COVID-19 ARDS drives a unique program of neutrophil maturation, activation, or effector functions that contributes to the severe pathogenesis of this pandemic illness, and whether this unique neutrophil response could be targeted therapeutically to supplement established therapies like dexamethasone. To address these questions, we used a combination of high-dimensional single cell analysis and *ex vivo* functional assays of neutrophils from patients with COVID-19 ARDS compared to non-COVID ARDS (bacterial pneumonia). We observed that COVID-19 programs a distinct landscape of neutrophil activation compared to non-COVID ARDS, and that neutrophil from COVID-19 patients are uniquely primed to produce NETs. This functional priming was an intrinsically programmed response during SARS-CoV-2 infection, as it could not be induced in healthy neutrophils by incubation with the plasma of COVID-19 patients, and neutrophils remained competent to respond to secondary bacterial challenges. Consistent with previous reports, we observed a trend towards worse clinical outcomes in patients with high levels of NETs production, but surprisingly we found that this was not modified in patients receiving dexamethasone treatment. Together, these data reveal a unique pathological program of neutrophil priming in COVID-19 that escapes conventional therapy, and therefore may represent a promising target for adjunctive treatments that provide synergistic benefits in the fight against severe COVID-19.

## RESULTS

### The systemic immune response in COVID-19 ARDS is dominated by a unique neutrophil landscape and is distinct from non-COVID ARDS

It is now well established that SARS-CoV-2 induces a unique systemic immune response that is central to the pathogenesis of severe COVID-19, including a putative role for neutrophils in the progression of disease during SARS-CoV-2 infection. To systematically examine the contribution of neutrophils to systemic immune dysregulation in severe COVID-19, we investigated the cellular immune landscape in the bloodstream of patients with ARDS caused by COVID-19 pneumonia or bacterial pneumonia (hereafter, non-COVID ARDS). Patients were balanced with respect to age, sex, illness severity, and treatments including mechanical ventilation (Table 1). Patients were enrolled prospectively, and blood specimens collected on admission (day 1) and again on day 7 in survivors who remained in the ICU. We performed high-dimensional single cell analysis using mass cytometry on whole blood from patients with COVID-19 ARDS compared to non-COVID ARDS. Neutrophils were the dominant immune cell in the blood of all patients, with the total quantity of neutrophils being higher in patients with non-COVID ARDS (Fig 1A). Comparatively small differences were seen in other immune cell populations such as classical monocytes, myeloid dendritic cells, and activated B-cells, with no significant differences found in other major cell compartments (Fig 1A). In addition to quantitative difference, we next investigated whether the neutrophils differed phenotypically between COVID-19 and non-COVID ARDS patients. Dimensionality reduction using tSNE revealed marked changes in the clustering of neutrophils between patients with COVID-19 and non-COVID ARDS, whereas clusters of other major cell populations (T-cells, B-cells, monocytes, etc.) were largely stable (Fig 1B). To further investigate this difference in the neutrophil compartment, we analyzed expression levels of 23 neutrophil-relevant surface and intracellular markers on gated neutrophils, and found significant differences driven by augmented expression of CD11b, CD66b, CD11a, and reduced CD62L, and CD107A on neutrophils from patients with COVID-19 ARDS, consistent with a more activated phenotype (Fig 1C and D). Together these data demonstrate a distinct neutrophil landscape in COVID-19 ARDS that is a dominant aspect of the systemic immune response to SARS-CoV-2.

**Table 1.** Study Patient Characteristics.

| **Characteristics** | **COVID-19 ARDS**<br>**(n=21)** | **Non-COVID ARDS**<br>**(n=10)** |
| --- | --- | --- |
| **Demographics** |  |  |
| Age - median (range) | 58 (36 - 84) | 56 (37 - 77) |
| Male sex - n (%) | 13 (61.9%) | 7 (70%) |
| **Cause of ARDS** |  |  |
| SARS-CoV-2 | 21 (100%) | 0 (0%) |
| Bacterial pneumonia (culture positive) | 0 (0%) | 8 (80%) |
| Bacterial pneumonia (culture negative) | 0 (0%) | 2 (20%) |
| **Clinical Characteristics** |  |  |
| Admission SOFA Score - median (range) | 9 (2 - 13) | 11 (5 - 15) |
| Admission P<sub>a</sub>O<sub>2</sub>/F<sub>i</sub>O<sub>2</sub> ratio - median (range) | 166 (72 - 285) | 185 (127 - 290) |
| Day 3 P<sub>a</sub>O<sub>2</sub>/F<sub>i</sub>O<sub>2</sub> ratio - median (range) | 190 (66 - 400) | 212 (157 - 380) |
| **Therapies** |  |  |
| Invasive mechanical ventilation - n (%) | 19 (90.5%) | 10 (100%) |
| Dexamethasone - n (%) | 12 (57.1) | 0 (0%) |
| Tocilizumab - n (%) | 0 (0%) | 0 (0%) |
| Remdesivir - n (%) | 1 (4.7%) | 0 (0%) |
| Hydroxychloroquine - n (%) | 2 (9.5%) | 1 (10%) |
| Antibiotics on admission - n (%) | 21 (100%) | 10 (100%) |
| **Outcomes** |  |  |
| Duration of mechanical ventilation - median days (range) | 11 (0 - 52) | 7 (3 - 44) |
| Duration of hospitalization - median days (range) | 34 (6 - 193) | 49 (9 - 135) |
| 90 day mortality - n (%) | 4 (19.0%) | 1 (10%) |

**Figure 1.**
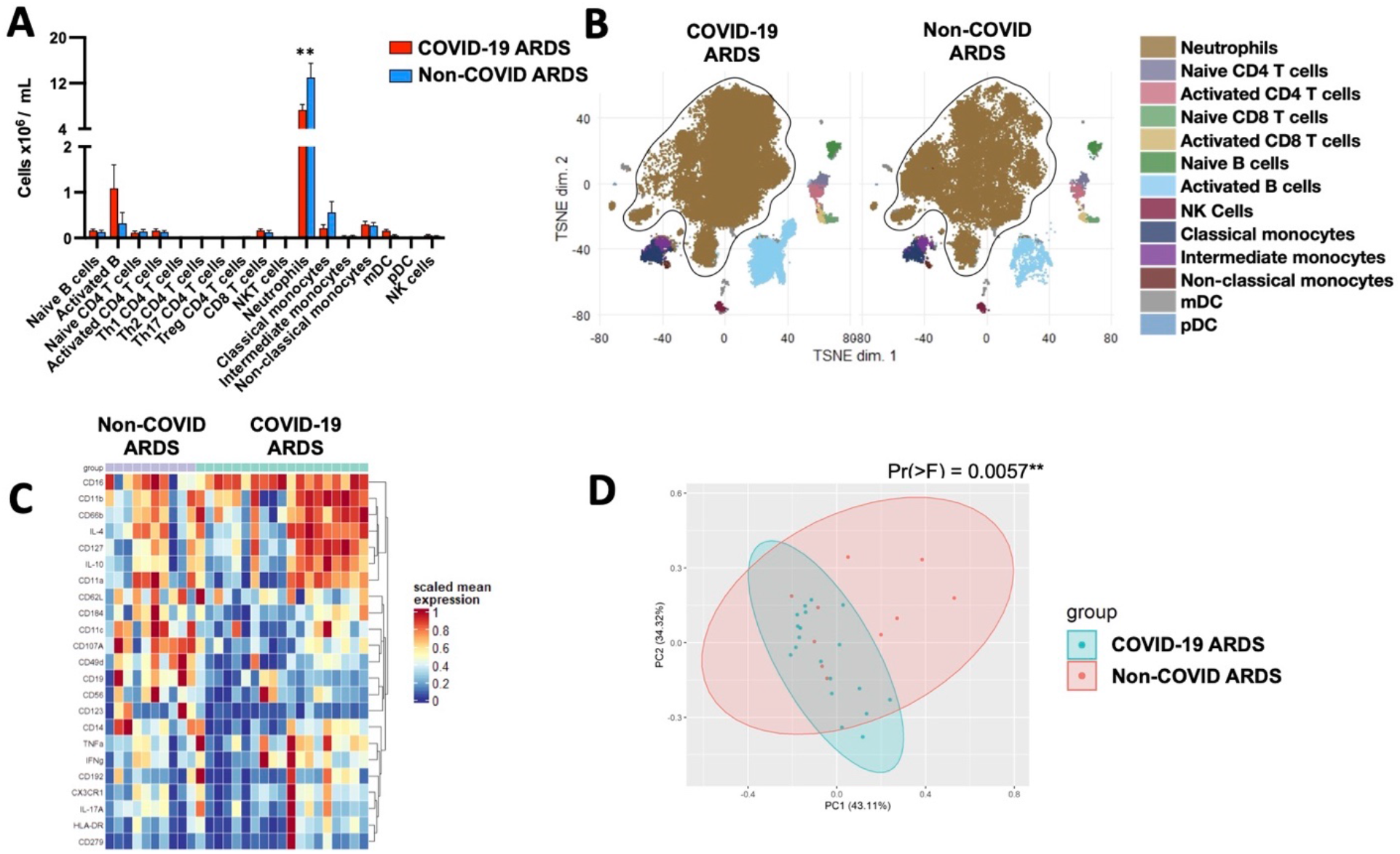
A distinct neutrophil landscape dominates the systemic immune response in COVID-19 ARDS. (A) Abundance of immune cell populations in the blood of patients with COVID-19 ARDS compared to non-COVID ARDS on day 1 of ICU admission. Data are mean +/- SEM. (B) Dimensionality reduction using tSNE of single cell mass cytometry data demonstrating clustering of major immune cell population in the blood of patients on day 1 of ICU admission. (C) Expression levels of selected neutrophil surface and intracellular markers, and (D) principal component analysis of neutrophils on day 1 of ICU admission in patients with COVID - 19 ARDS and Non-COVID ARDS. **p<0.01.

To further resolve the differences between COVID-19 and non-COVID neutrophil responses, we performed unsupervised clustering of neutrophil single cell events with FlowSOM, and identified 8 distinct clusters of neutrophils (Fig 2A). Comparing our two patient populations, we identified stark differences in the presence and abundance of distinct neutrophil clusters between COVID-19 and non-COVID ARDS patients (Fig 2A-C). Again, we observed dominance by mature and activated neutrophil clusters in all patients with COVID-19, whereas the neutrophil compartment in patients with non-COVID ARDS was more heterogeneous, with an overall predominance of immature neutrophil populations (Fig 2C-D). Collectively, these data indicate that patients with COVID-19 ARDS display a unique systemic immune response that is dominated by a distinct neutrophil landscape compared to non-COVID ARDS, and is driven by a shift towards a more mature and activated neutrophil phenotype.

**Figure 2.**
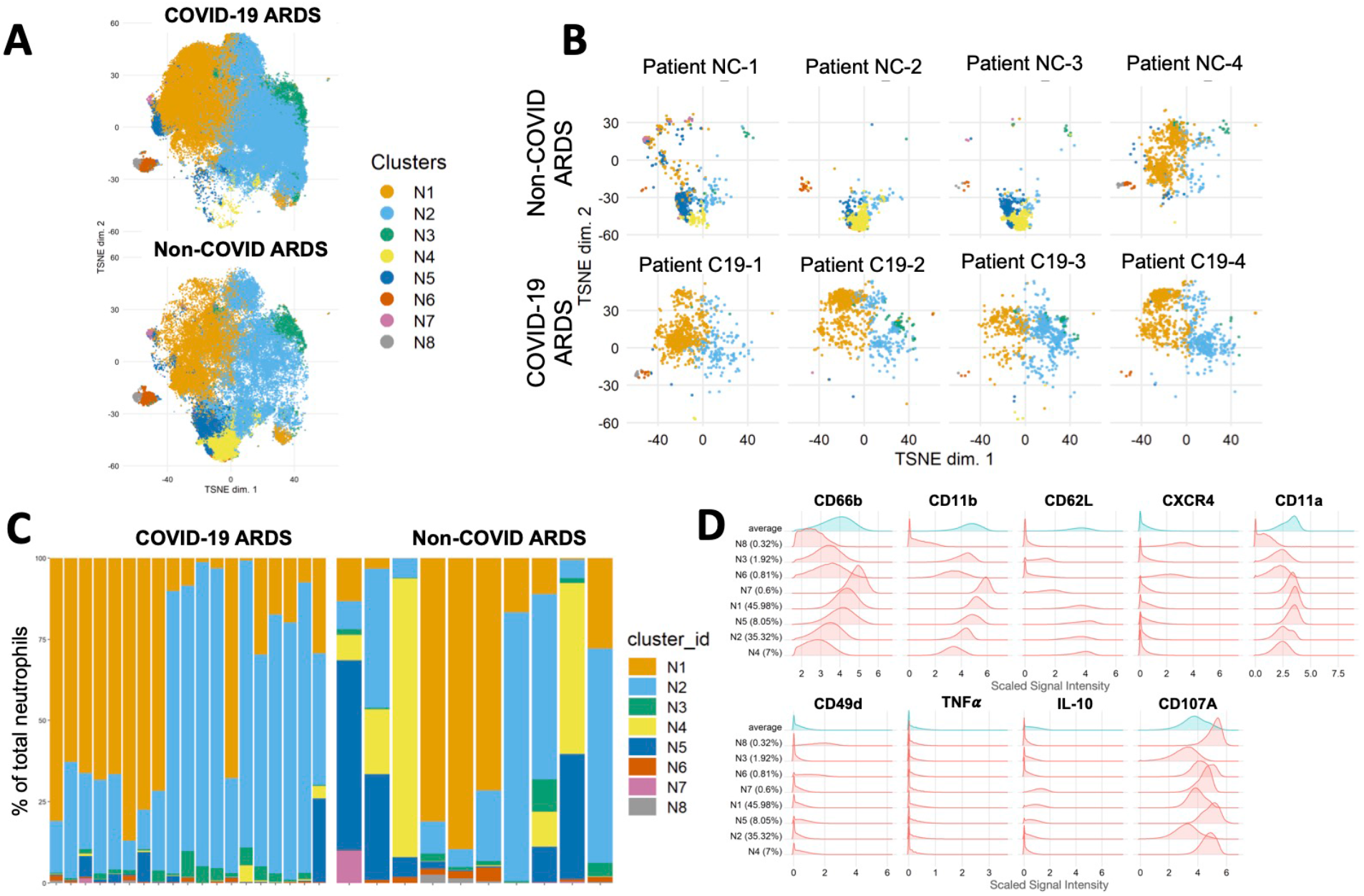
A distinct neutrophil landscape in COVID-19 ARDS versus non-COVID ARDS. Mass cytometry analysis of neutrophils showing (A) unsupervised clustering of neutrophil single cell events using FLOWSOM in COVID-19 ARDS (n=19) and non-COVID ARDS (n=10) patient samples, and (B) representative clustering of neutrophils selected individual patients with COVID-19 ARDS and non-COVID ARDS. (C) Relative abundance of neutrophils clusters in individual patient samples, and (D) histogram plots demonstrating expression of key surface and intracellular markers that differentiate the identified neutrophil clusters.

### The unique neutrophil landscape in COVID-19 ARDS is associated with functional priming

Given the distinct phenotypic landscape of neutrophils observed in COVID-19 ARDS, we next sought to determine whether neutrophils in COVID-19 ARDS display unique effector functionality. We performed *ex vivo* live-cell functional analyses on neutrophils isolated from patients with COVID-19 versus non-COVID-19 ARDS to quantitatively assess key effector function including the release of neutrophil extracellular traps (NETs) and reactive oxygen species. To understand the intrinsic functional activity of neutrophils in response to COVID-19 or non-COVID-19 ARDS, we first assessed the production of NETs and ROS in otherwise unstimulated cells. Surprisingly, neutrophils from patients with COVID-19 ARDS on day 1 of ICU admission displayed significantly higher basal NETs release compared to neutrophils from patients with non-COVID ARDS (Fig 3A-B). As expected, neutrophils from healthy volunteers did not release NETs under basal conditions (Fig 3A-B). This augmented NETs release in COVID-19 neutrophils was sustained over time, with a notable trend towards increased production of NETs on day 7 of illness in patients with COVID-19 but not non-COVID-19 ARDS (Fig 3C). Neutrophils from all groups were found to release similar levels of NETs upon stimulation with phorbol myristate acetate-13 (PMA) suggesting that the capacity to respond to maximal activation was not different (Supp. Fig. 1). In contrast to NETs release, ROS production by neutrophils was not different between COVID-19 and non-COVID ARDS either at admission or after 7 days in the ICU (Fig. 3D-F). Together, these data indicate that the distinct neutrophil landscape in COVID-19 ARDS is marked by functional priming of specific effector mechanisms including neutrophil extracellular trap release.

**Figure 3.**
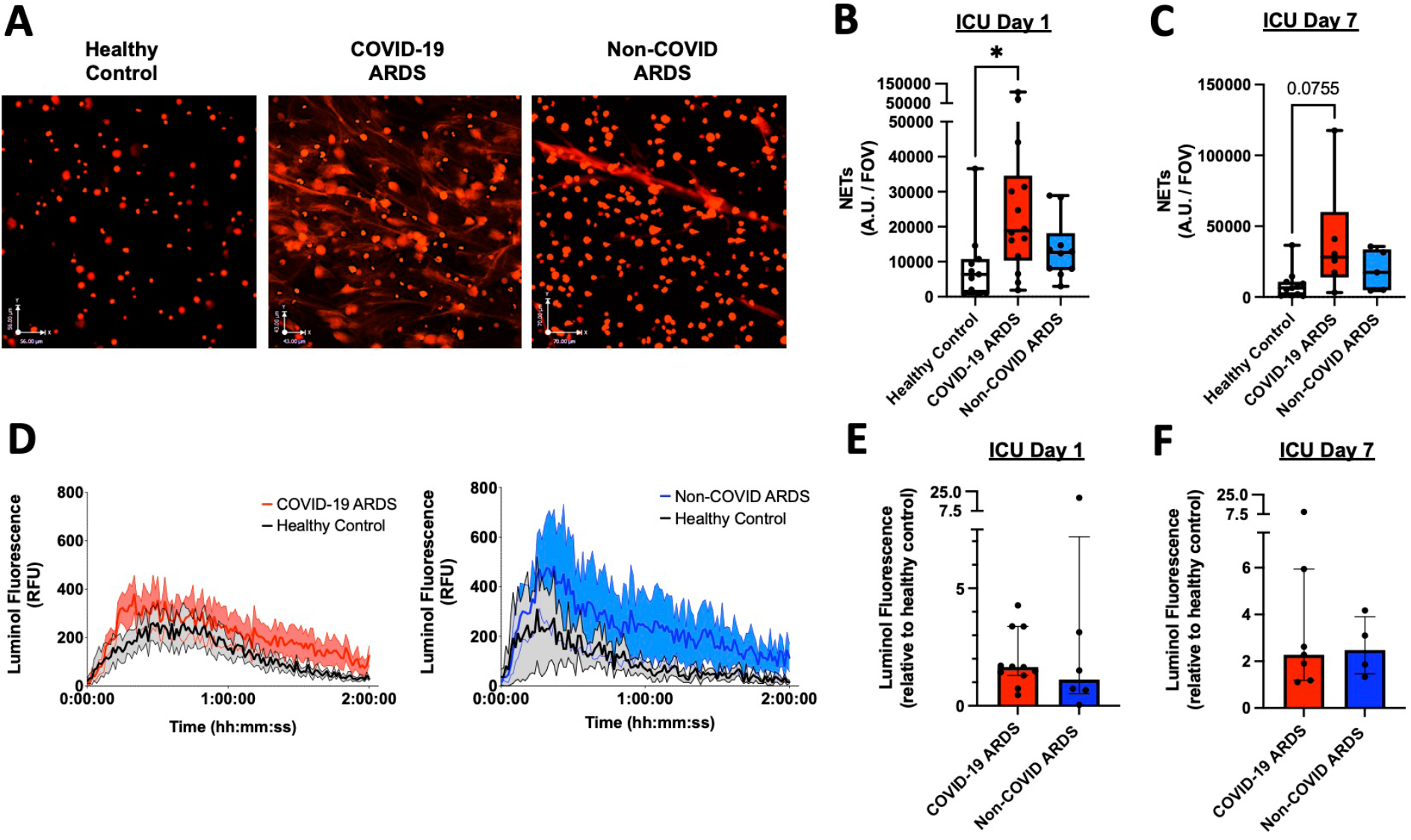
Neutrophils in COVID-19 ARDS are uniquely primed to produce neutrophil extracellular traps. (A) Representative images of NET release from healthy control (left), COVID-19 ARDS patient (center) and non-COVID ARDS patient (right) neutrophils. Scale bars represents 75 µm. (B-C) Quantitation of area per field of view covered by NETs from neutrophils in patients with COVID-19 and non-COVID ARDS on day 1 (B) and at day 7 (C) after ICU admission and healthy controls. Data are individual patients, with median and range. (D-F) Reactive oxygen species (ROS) production by neutrophils detected by luminol fluorescence assay shown as (D) relative fluorescence units (RFU) over time and (E-F) AUC of luminol fluorescence of patient neutrophils relative to healthy control neutrophils on day 1 (E) and day 7 (F) following ICU admission. Data are individual patients, with median and interquartile range.

### Functional priming of neutrophils in COVID-19 is not induced by circulating inflammatory mediators

Plasma biomarker analysis in various cohorts of patients with severe COVID-19 have described a circulating “cytokine storm” that is distinct from other causes of sepsis and ARDS (16). Previous work by us and others (17, 18) has shown that circulating inflammatory mediators in patients with septic shock induce neutrophil hyper-production of effector mechanisms, including the release of neutrophil extracellular traps. To determine whether the functional priming and activation of neutrophils observed in COVID-19 ARDS was a response to the unique circulating cytokine storm in these patients, we assessed the ability of patient plasma to induce NETs release by autologous neutrophils isolated from healthy volunteers. Consistent with findings from previous studies, plasma from patients with non-COVID ARDS induced robust NETs release by healthy donor neutrophils (Fig 4A). In stark contrast, plasma from patients with COVID-19 failed to induce NETs release (Fig 4A). There was also no induction of ROS generation in response to plasma from patients with COVID-19 nor non-COVID ARDS (Fig 4B-C). These data demonstrate that functional priming and augmented NETs production of COVID-19 neutrophils is not a by-product of a unique circulating inflammatory milieu (“cytokine storm”) in patients with COVID-19.

**Figure 4.**
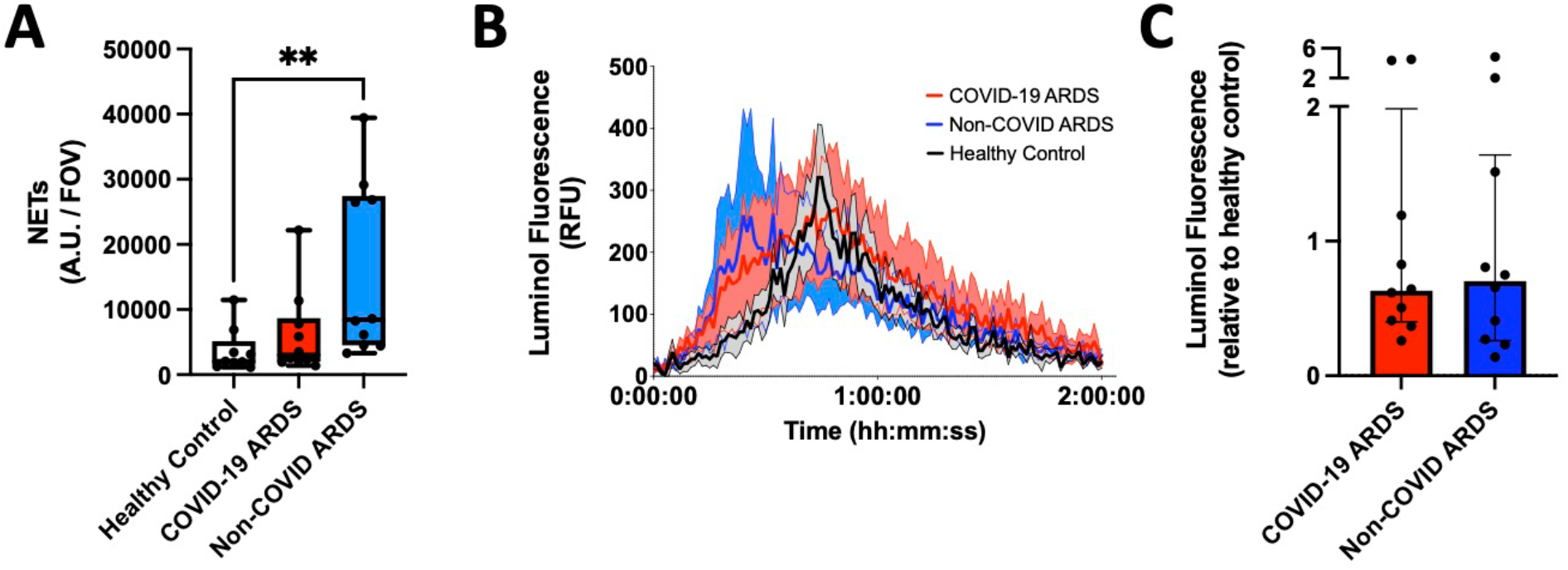
Neutrophil priming is not induced by inflammatory mediators in plasma. (A) *Ex vivo* imaging of NETs release quantified by NETs area per field of view from neutrophils from healthy volunteers incubated with plasma of healthy controls (n=9), COVID-19 ARDS (n=10) and non-COVID ARDS patients (n=10). Data are individual patients, with median and range. (B-C) Reactive oxygen species (ROS) production by neutrophils detected by luminol fluorescence assay shown by (B) relative fluorescence units (RFU) and (C) AUC of luminol fluorescence following stimulation with patient plasma relative to healthy control plasma. Data are individual patients, with median and interquartile range. **p<0.01.

### Priming of neutrophils in COVID-19 does not alter the response to secondary bacterial challenge

In addition to their role as pathogenic effector cells in ARDS, neutrophil dysfunction in critically ill patients also contributes to other complication of ARDS, including high susceptibility to secondary bacterial infections (including ventilator-associated pneumonia) (19). Therefore, we sought to determine if the distinct neutrophil program in COVID-19 ARDS may drive differential susceptibility to secondary bacterial infections compared to non-COVID ARDS. To test this hypothesis, neutrophils were challenged *ex vivo* with the common nosocomial bacterial pathogen *Staphylococcus aureus* (MRSA, MW2 strain) to determine their ability to mount anti-bacterial effector mechanisms (phagocytosis, NETs production, ROS production) in response to secondary bacterial challenge. Live cell imaging of neutrophils isolated from patients with COVID-19 and non-COVID ARDS demonstrated robust capture and phagocytosis of fluorescent *S. aureus* (GFP^+^) in both patient groups on admission and after 7 days in ICU, and was not different from healthy donor neutrophils (Fig 5A and 5B). Furthermore, production of NETs (Fig 5C-D) and ROS (Figure 5E-F) in response to *S. aureus* stimulation was also similar between COVID-19 and non-COVID ARDS patients. These equivalent effector responses to secondary bacterial challenge is consistent with the finding that nosocomial infections occurred in similar proportions in each patient group, as shown by equivalent infection-free survival between patient populations (Fig 5G). This is consistent with published reports of nosocomial infections in severe COVID-19 (20). Together, these results suggest that neutrophils retain their competence to respond to secondary bacterial pathogens during COVID-19 ARDS, consistent with the observation that difference in clinical outcomes with non-COVID critical ARDS are not driven by differential susceptibility to nosocomial infections (20).

**Figure 5.**
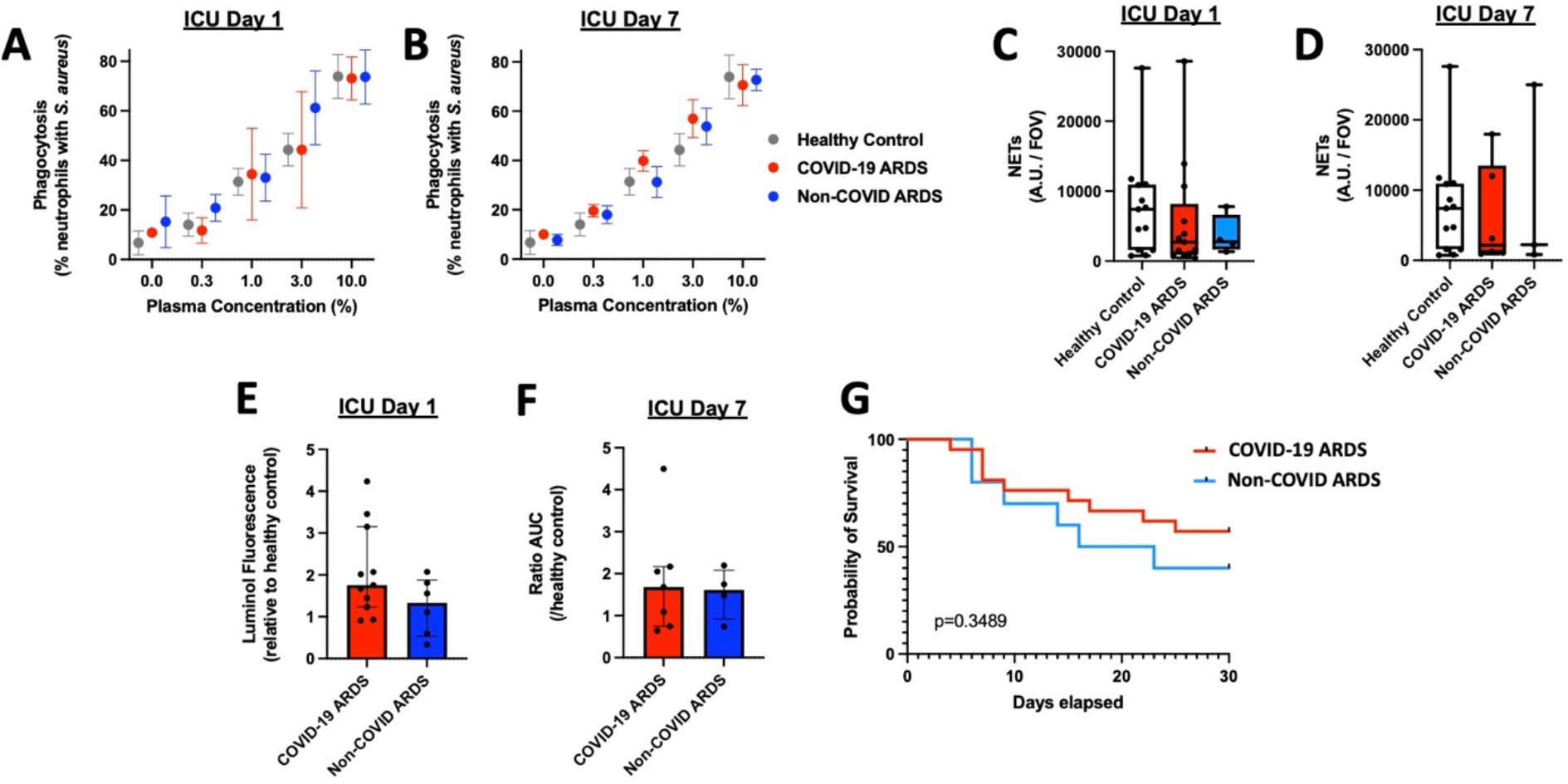
Neutrophils in COVID-19 and non-COVID ARDS remain functionally competent to respond to secondary bacterial challenge. (A-B) Quantitative assessment of phagocytosis of bacteria (GFP-expressing *S. aureus*) by neutrophils using flow cytometry on (A) days 1 and (B) day 7 of ICU admission, as well as healthy controls. Data are percentage of neutrophils containing GFP^+^ *S. aureus*, expressed as mean +/- SEM (Day 1: COVID-19 ARDS n=5, non-COVID ARDS n=3, healthy controls n=5; Day 7: COVID-19 n=4, non-COVID n=3, healthy controls n=5). (C-D) Quantification of NETs production after stimulation with *S. aureus* by neutrophils from patients and healthy controls on (C) day 1 (D) and day 7 of ICU admission. Data are individual patients, with median and range. (E-F) ROS production by neutrophils detected by luminol fluorescence assay, shown by AUC of luminol fluorescence following stimulation with *S. aureus*. Data are individual patients, with median and interquartile range. (G) Probability of nosocomial infection-free survival of patients with COVID-19 ARDS (n=21) and non-COVID ARDS (n=10) patients from ICU admission (day 0) to day 30.

### Pathological neutrophil priming in COVID-19 may escape established therapies, revealing opportunities for targeted therapeutic adjuncts

Emerging data implicates hyperactive NETs as an important mediator of pulmonary vascular and parenchymal injury in COVID-19, with recent studies demonstrating an association between surrogate markers of NETs in the circulation and disease severity (21, 22). Given that augmented NETs release is a key feature of the functionally distinct neutrophil program observed in our COVID-19 patients, we analyzed the impact of NETs production on clinical outcomes. Although our sample size was insufficient to achieve statistical significance, a clear trend was observed with 100% 90-day survival in low NETs producers (neutrophil NETs release <cohort median), compared to only 60% 90-day survival in high NETs producers (neutrophil NETs release >median of cohort) (Fig 6A). Together with emerging published data, these findings suggest that NETs may be an important pathogenic mediator in COVID-19 and may be amenable to therapeutic targeting (23). Therefore, we aimed to determine whether the clinical efficacy of dexamethasone therapy may be related to its ability to modulate pathological neutrophil priming in COVID-19. We analyzed NETs production, surface and intracellular marker expression, and single cell cluster profiles of neutrophils from COVID-19 patients who were treated with dexamethasone compared to patients who did not receive dexamethasone. Surprisingly, we observed no significant difference in NETs production nor the phenotypic landscape of circulating neutrophils after treatment with dexamethasone, either early (day 1) or later (day 7) during ICU admission (Fig 6B-E, Supplementary Fig 2). Given that pathological neutrophil priming and NETs production are not modified by dexamethasone treatment, these data suggest that adjunctive therapies (eg. NETs targeted therapies) may yield additive benefits to dexamethasone and represents an important avenue for further therapeutic development in the fight against COVID-19 ARDS.

**Figure 6.**
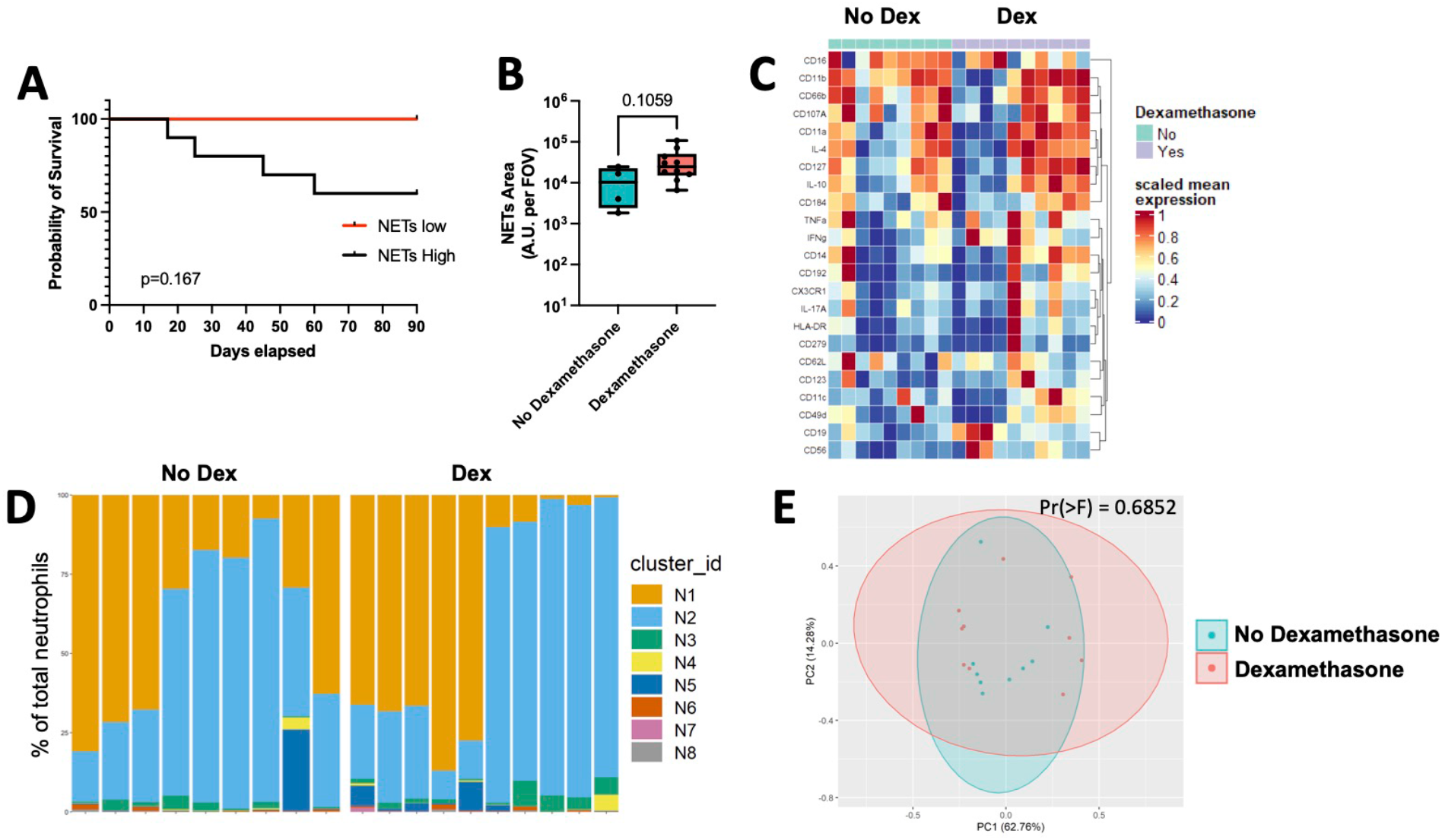
Pathological neutrophil priming in COVID-19 escapes treatment with dexamethasone. (A) Probably of survival following admission to ICU (day 0) to 90 days in patients with COVID-19 whose neutrophils produced high (>cohort median) or low (<cohort median) levels of NETs. (B) Quantification of NETs production (area per field of covered by NETs) by neutrophils from patients with COVID-19 ARDS who received dexamethasone treatment versus those who did not. Data are individual patients on ICU day 1, with median and range. (C-E) Mass cytometry analysis of neutrophils from patients with COVID-19 ARDS (ICU day 1) who received dexamethasone treatment versus those who did not, showing (C) expression levels of selected neutrophil surface and intracellular markers, (D) relative abundance of neutrophils clusters in individual patient samples determined by FlowSOM analysis, and (E) principle component analysis of neutrophils.

## DISCUSSION

Our results add to the growing literature on the importance of neutrophils in the immunopathogenesis of ARDS, including severe COVID-19. It is now well established that patients with SARS-CoV-2 infection display widespread alterations in the myeloid compartment that contribute to disease severity (24, 25). Using high-dimensional single cell analysis of the entire immune landscape in whole blood, we further this understanding by demonstrating that the systemic immune response in COVID-19 ARDS is dominated by a unique neutrophil compartment characterized by mature and active neutrophil populations. This was in stark contrast to the neutrophil response during non-COVID ARDS, which was much more heterogeneous and dominated by immature neutrophil populations. Further dissecting the functional implications of this unique neutrophil program in COVID-19, we observed that neutrophils from patients with COVID-19 ARDS were functionally primed to release neutrophil extracellular traps, and that high-NETs producers were at greater risk of adverse clinical outcomes, consistent with previous reports (21–23). Surprisingly, this functional priming was not attenuated in patients treated with dexamethasone therapy, thereby revealing this pathological neutrophil program as a potential target for adjunctive therapies to combat COVID-19 ARDS.

We found that neutrophils are functionally primed in COVID-19 ARDS to produce NETs, and that this priming was intrinsically programmed during SARS-CoV-2 infection and could not be induced in healthy neutrophils in response to plasma from patients with COVID-19 ARDS. Again, this differed from non-COVID ARDS, as plasma from these patients induced robust NETs production by healthy donor neutrophils, consistent with previous studies that have shown NETs production in response to plasma from patients with bacterial sepsis and ARDS (17, 18). The “cytokine storm” of severe SARS-CoV-2 infection has been the focus of extensive investigation to understand the systemic immune drivers of COVID-19. A systematic review and meta-analysis of published cytokine data from 25 studies (totalling 1245 patients) found that substantial differences existed in the levels of core pro- and anti-inflammatory cytokines between those with COVID-19 ARDS and non-COVID-19 ARDS, revealing that levels of key mediators like IL-6 and TNFα were often much lower in COVID-19 ARDS (16). Together with our findings, these data collectively support a unique systemic milieu of inflammatory mediators in COVID-19 compared to non-COVID ARDS that have differential effects on neutrophil function.

Amongst the arsenal of neutrophil effector mechanisms, NETs are emerging as important pathological mediators of ARDS in COVID-19 (23). Histopathological analysis of lung tissue from autopsies of patients who have succumbed to COVID-19 have consistently shown that neutrophil infiltration in the lungs is closely associated with NETs release, as well as downstream sequelae including microvascular immunothrombosis (11–14). Quantitative assessment of surrogate markers of NETs in the bloodstream including DNA-MPO complexes and citrullinated histone-3 have suggested that higher NETs production is directly correlated with disease severity, and may even predict patients who will progress from mild to severe disease (21, 22). This is consistent with our observation that neutrophils from patients with COVID-19 are primed to produce NETs, and that there was a trend towards higher mortality in patients who produced high levels of NETs. The exact mechanisms that propagate NETs production during severe COVID-19 remain an active area of investigation, and may include direct stimulation and/or infection of neutrophils by SARS-CoV-2, as well as autoantibody-mediated stabilization of NETs structures *in vivo* (12, 26). Our data suggest that the majority of neutrophils in patients with severe COVID-19 display a mature and functionally active phenotype and are intrinsically primed to release NETs, which differs markedly from neutrophils in non-COVID ARDS, further underpinning the potential importance of this stereotyped immune response to the unique pathogenesis of COVID-19.

The findings of this study lend further support to the potential therapeutic utility of targeting pathological neutrophil mechanisms, including NETs, for the treatment of COVID-19 ARDS (23). In particular, we found that dexamethasone treatment did not modulate the functional landscape of neutrophils nor attenuate the production of NETs. This is in line with a recent pre-print publication suggesting that dexamethasone does not alter the NETs-related neutrophil proteome in patients with severe COVID-19 (27). Of note, our findings do not rule out the possibility that dexamethasone modulates other aspects of neutrophil development or activation *in vivo*, as has been suggested using transcriptomic analysis of the circulating neutrophil pool in severe COVID-19 (28). Together, these data suggest that neutrophil-mediated pathogenesis including augmented NETs production may escape the treatment effect of dexamethasone, and therefore represent a promising avenue for therapeutic adjuncts to supplement current treatment regimens.

Lastly, this study also uncovers a number of outstanding questions regarding the role of neutrophils in the pathogenesis of COVID-19 and ARDS. First, our study focuses on the immune landscape in the bloodstream compartment, but understanding the implications of our findings within the lung microenvironment may reveal important features of tissue-specific neutrophil responses in COVID-19. A recent study of the single cell landscape of lung immunity in SARS-CoV-2 infection, including spatially-resolved profiling with imaging mass cytometry, also observed a distinct pattern of neutrophil infiltration in the lungs of patients with severe COVID-19 compared to non-COVID pneumonia (9). However, much remains to be learned about whether there is selective recruitment of specific neutrophil populations to the lungs in COVID-19 (eg. NETs-producing neutrophils), as well as defining the molecular mechanisms of neutrophil recruitment within the pulmonary microvasculature. Indeed, it was recently reported that neutrophil recruitment to the lungs in ARDS utilizes a unique adhesion molecule (DPEP-1), which is now being targeting in clinical trials for moderate to severe COVID-19 (ClinicalTrials.gov NCT04402957) (29). In addition, outstanding questions remain about the molecular mechanisms that program the systemic neutrophil landscape in COVID-19, as well as further defining the mechanisms that elicit pathological NETs release within the lung tissues. Finally, our modest sample size was necessary to enable deep profiling of neutrophil phenotypes and functions, but limits our ability to address their impact on clinical outcomes. Although we did observe an association between neutrophil priming and high NETs production with mortality in COVID-19 ARDS, a larger study will be required to confirm the impact of our findings on adverse clinical outcomes in severe COVID-19 (degree of hypoxemia, illness severity, multi-organ dysfunction, thrombotic complications, and death). However, the demonstration of a functionally distinct neutrophil landscape in COVID-19 that escapes conventional therapy with dexamethasone provides further support for the ongoing efforts to develop neutrophil/NETs-targeted therapies to treat COVID-19 ARDS.

## MATERIALS AND METHODS

### Study design

Written informed consent was obtained from all study participants or most appropriate surrogate decision maker for patients who were unable to provide consent. Between April 1 2020 and March 30 2021, consecutive patients were screened for the following inclusion criteria: adult patients (>18 years of age) with an index admission to one of four multi-system intensive care units in Calgary, AB, Canada, with a diagnosis of ARDS based on the Berlin Criteria (30), associated with a diagnosis of SARS-CoV-2 infection (based on clinical laboratory qPCR assay from nasopharyngeal swab or endotracheal tube aspirate) or bacterial pneumonia (based on standard clinical and microbiologic criteria). Exclusion criteria included pre-existing immunocompromised state (immunomodulatory therapy, chemotherapy, HIV infection, other congenital or acquired immunodefiency), re-admission to ICU or prior study enrollment, ARDS due to another cause, goals of care that excluded life-support interventions, or moribund patients not expected to survive >72 hours. Blood samples were collected from enrolled patients with COVID-19 (n=21) and non-COVID (n=10) ARDS after admission to ICU on days 1 and 7 (for those who survived and remained in ICU). Blood samples were also collected from healthy volunteers for use as controls. This study was approved by the conjoint health research ethics board of the University of Calgary and Alberta Health Services (REB18-1294).

### Time-of-flight mass cytometry and analysis

Whole blood samples used for mass cytometry analysis were cryopreserved in PROT1 proteomic stabilizer (SmartTube) at a ratio of 1:1.4 and stored at -80ºC to enable batched analysis of patient samples as previously described (31). Samples were thawed at room temperature, followed by RBC lysis using PROT1 RBC lysis buffer (SmartTube), and white blood cells were washed in cell staining medium (PBS with 1% BSA) followed by labelling with a custom metal-conjugated antibody panel (supplementary table 1). First, cells were incubated with metal-conjugated surface antibodies, followed by fixation and permeabilized (BD Cytofix-Cytoperm), and incubation with intracellular antibodies. Finally, cells were incubated overnight in a solution containin Cell-ID iridium intercalator (Fluidigm), 0.3% saponin, and 1.6% paraformaldehyde in PBS. Cells were then mixed with EQ Four Element Calibration Beads (Fluidigm), and acquired on a Helios CyTOFII mass cytometer (DVS). Mass cytometry data was normalized using the internal Helios CyTOFII bead-based normalization software (DVS).

For mass cytometry data analysis, FCS files were imported into Cytobank (Cytobank.org) for data visualization and manual gating. Single-cell events were then exported from CytoBank as FCS files and loaded into the CATALYST package (32) in R (R Development Core Team). For neutrophil clustering and analysis, CD45^+^CD66b^+^ neutrophils were analyzed by FlowSOM (33) and a Consensus Clustering method (34) was employed on all neutrophil single-cell events within CATALYST based on the expression of neutrophil markers (see Fig 1C). To determine the optimal number of metaclusters, we compared the change in the area under the curve of the CDF plot for each additional metacluster added (*delta_area* function; CATALYST). Using this method, we identified 11 neutrophil metaclusters. Very rare metaclusters (represented less than 0.5% of total neutrophils) were excluded from the analysis, leaving 8 metaclusters in the analysis. To visualize the neutrophil metacluster landscape, tSNE dimensionality reduction was performed on 2,500 randomly selected events from each sample using a perplexity of 70 for 5000 iterations. Data visualization was generated using the built-in functions of CATALYST and the ggplot2 package in R.

### Plasma and neutrophil isolation

Blood samples collected in heparinized tubes were centrifuged at 450 g and pelleted blood cells were resuspended in phosphate-buffered saline (PBS), and neutrophils were isolated using a two density histopaque gradient (1.119 g/mL and 1.077 g/mL; Sigma cat. 11191 and 10771, respectively) with centrifugation at 400 g for 20 minutes at 20 °C without braking. The layer of neutrophils (second layer) was collected, red blood cells were lysed, and isolated neutrophils were resuspended in RPMI containing 0.05% of human serum albumin at a final concentration of 1×10^7^ neutrophils/mL. Neutrophil viability was >98% as determine by trypan blue staining. Of note, not all patient samples yielded sufficient quantities of neutrophils to conduct every *ex vivo* neutrophil assays, but every effort was made to include as many patients as possible in all assays.

### NETs release assay

Isolated neutrophils (5 × 10^4^) were seeded into sterile 96-well optical plates (Falcon) and allowed to settle for 5 minutes at room temperature. The cells were incubated under the following conditions: unstimulated, 0.1 µM PMA, or 1 × 10^6^ CFU of *S*.*aureus* MW2 for 3 hours at 37°C with 5% CO_2_. To assess NET release in response to patient plasma, the isolated neutrophils from healthy volunteers were incubated with 5% patient plasma containing 5mM EDTA for 3 hours at 37°C with 5% CO_2._ Cells were then fixed with 2% paraformaldehyde (PFA) overnight, and then stained with Sytox Orange (Thermofisher) to visualize extracellular DNA NETs and imaged on an inverted spinning disk confocal microscope with a 10X objective. The quantity of NETs release was determined by measuring that visualized area covered by extracellular DNA NETs per field of view.

### Reactive oxygen species production assay

Isolated neutrophils (1×10^6^) were incubated with luminol (50uM) in the presence of superoxide dismutase (75 ug/ml), catalase (2000 U/ml), and horsedarish peroxidase (20 U/ml), in an opaque 96 well plate. In some experiments, neutrophils were co-incubated with *S. aureus* MW2 (1 × 10^6^ CFU). Chemiluminescence was read using Spectramax i3x instrument (Molecular Devices, San Jose, Ca, USA) every 1 minute for 120 minutes.

### Phagocytosis assay

Methicillin-resistant *S*.*aureus* MW2, constitutively expressing EGFP, was grown o/n in Brain Heart Infusion broth supplemented with 10 μg/mL chloramphenicol. On the day of the experiment, the strain was sub-cultured to late log phase (OD_660nm_ 1.0), washed, and resuspended in RPMI-containing 0.05% human serum albumin. Subsequently, 2×10^5^ isolated neutrophils were mixed with 1×10^6^ bacteria (MOI=5) in the presence of various concentrations of normal human pooled serum (15 donors) in a final volume of 200 μL of RPMI containing 0.05% HAS, and incubated at 37°C with agitation for 15 min. The cells were then fixed with 4% paraformaldehyde and phagocytosis was measured using flow cytometry (FACS Canto). Neutrophils were gated based on their forward and side scatter profiles, and bacterial phagocytosis was measured by as the percentage of neutrophils positive for GFP^+^ bacteria using FlowJo software (FloJo, BD Biosciences).

### Statistics

Non-parametric data are represented as median +/- interquartile range, and analyzed using Mann-Whitney U test (when comparing 2 groups), or Kruskal-Wallis Test with post-hoc Dunn’s test for multiple comparisons (when comparing more than 2 groups). Parametric data are represented with mean +/- SEM, and analyzed using student’s T test (when comparing 2 groups), or one-way ANOVA with post-hoc Bonferroni correction for multiple comparisons when more than 2 groups were compared. Principal component analysis of neutrophil markers expression between patient groups was performed using the *procomp* function and VEGAN package in R, and plotted using ggplot2. PERMANOVA analysis of the Bray-Curtis distances between individual samples was used to compare PCA ordinations. Statistical analysis and graphic generation were performed using Graphpad Prism (GraphPad Software) and R (R Development Core Team).

## Data Availability

Data are available from the authors upon request.

## AUTHOR CONTRIBUTIONS

RP, FVSC, HS, BS and BM contributed to the design of the study, data acquisition and data analysis. JS and AZ made contributions to data acquisition and data analysis. ZS, BY, and BM contributed to patient enrollment, sample collection and processing, and clinical data acquisition. PK contributed to the design of the study and data analysis. RP, FVSC, JS, and BM wrote the manuscript, and all authors contributed to editing and approved the manuscript.

## ACKNOWLEDGMENTS

The authors thank the patients and families who agreed to participate in this study, and the many ICU nurses who aided in the collection of patient samples. We also thank Olesya Dmitrieva, and Cassidy Codan for assistance with screening, consenting, and enrolling patients. We also thank Dr. Anowara Islam and Dr. Karen Poon from the International Nicole Perkins Microbial Core Laboratory for the assistance with the CyTOF experiments. Funding was provided the Canadian Institutes of Health Research grant 202005VR3-172628 to PK and BM.

---

**Supplementary Figure 1.**
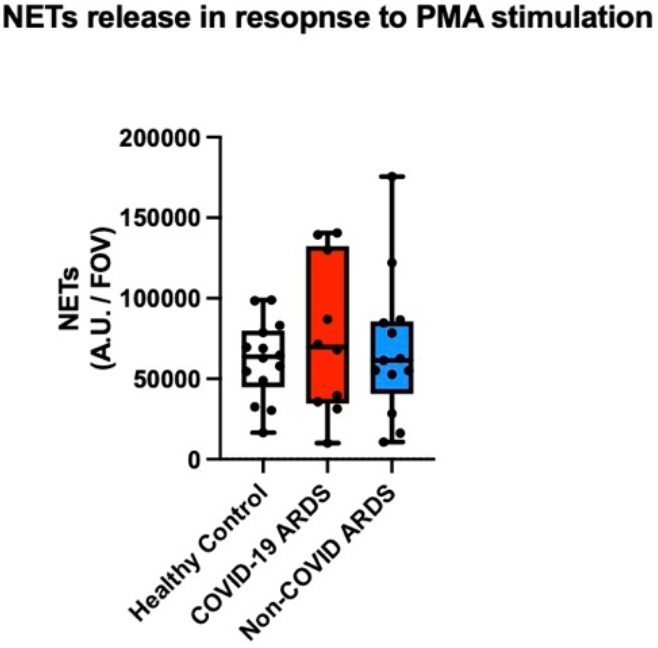
Neutrophils release similar levels of NETs upon stimulation with PMA. Quantification of NETs production (area per field of view covered by NETs) following stimulation with PMA by neutrophils from healthy controls (n= 14), COVID-19 ARDS (n= 10) and non-COVID ARDS (n= 13) following stimulation with PMA. Data are individual patients, with median and range.

**Supplementary Figure 2.**
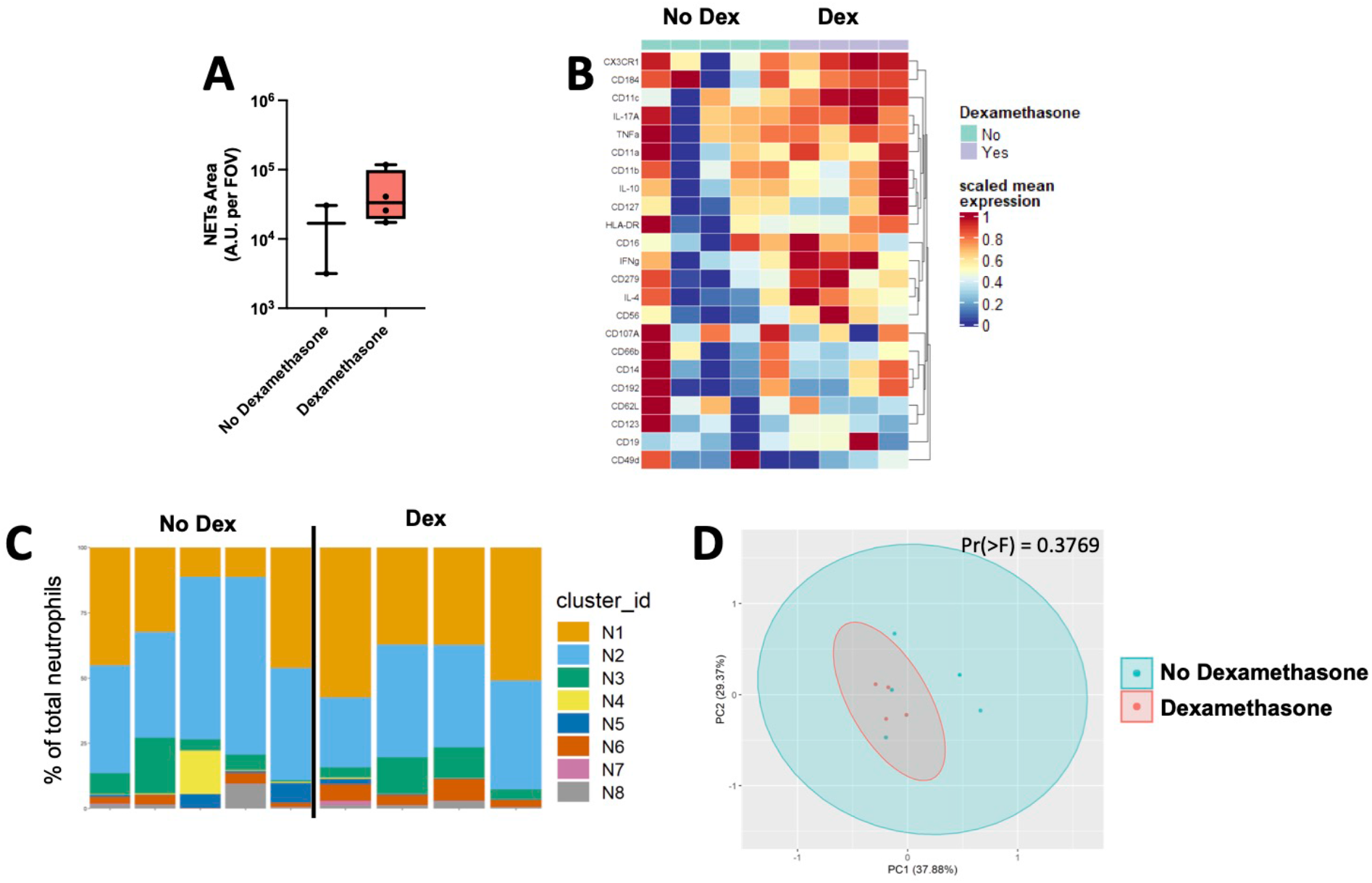
Figure 6. Pathological neutrophil priming in COVID-19 escapes treatment with dexamethasone on ICU day 7. (A) Quantification of NETs production (NETs area per field of view) by neutrophils from patients with COVID-19 ARDS who received dexamethasone treatment versus those who did not. Data are individual patients on ICU day 7 of ICU admission, with median and range. (B-D) Mass cytometry analysis of neutrophils from patients with COVID-19 ARDS (ICU day 7) who received dexamethasone treatment versus those who did not, demonstrating (B) expression levels of selected neutrophil surface and intracellular markers, (C) relative abundance of neutrophils clusters in individual patient samples determined by FlowSOM analysis, and (D) principle component analysis of neutrophils.

**Supplementary Table 1.** Mass cytometry antibody panel.

| Marker | Clone | Metal | Source |
| --- | --- | --- | --- |
| CD45 | HI30 | 89Y | Fluidigm |
| CD3 | UCHT1 | 141Pr | Fluidigm |
| CD19 | HIB19 | 142Nd | Fluidigm |
| CD107A | H4A3 | 143Nd | Biolegend* |
| CD4 | RPA-T4 | 145Nd | Fluidigm |
| CD8a | RPA-T8 | 146Nd | Fluidigm |
| CD11c | Bu15 | 147Sm | Fluidigm |
| IL-17A | BL168 | 148Nd | Fluidigm |
| CD56 | NCAM16.2 | 149Sm | Fluidigm |
| CD192 | M-T701 | 150Nd | Biolegend* |
| CD123 | 6H6 | 151Eu | Fluidigm |
| TNFα | Mab11 | 152Sm | Fluidigm |
| CD62L | DREG-56 | 153Eu | Fluidigm |
| CD49d | 9F10 | 154Sm | Biolegend* |
| CD45RA | HI100 | 155Gd | Fluidigm |
| CD184 | 12G5 | 156Gd | Fluidigm |
| IFNγ | B27 | 158Gd | Fluidigm |
| FoxP3 | 259D/C7 | 159Tb | Fluidigm |
| CD14 | M5E2 | 160Gd | Fluidigm |
| CD66b | 80H3 | 162Dy | Fluidigm |
| IL-4 | MP4-25D2 | 163Dy | Fluidigm |
| Granulysin | RB1 | 165Ho | Biolegend* |
| IL-10 | JES3-9D7 | 166Er | Fluidigm |
| CD11b | ICRF44 | 167Er | Fluidigm |
| CD127 | A019D5 | 168Er | Fluidigm |
| CD25 | 2A3 | 169Tm | Fluidigm |
| CD11a | HI111 | 170Er | Blolegend* |
| Granzyme B | GB11 | 171Yb | Fluidigm |
| CX3CR1 | 2A9-1 | 172Yb | Fluidigm |
| HLA-DR | L243 | 173Yb | Fluidigm |
| CD279 | EH12.2H7 | 174Yb | Fluidigm |
| Perforin | BD48 | 175Lu | Fluidigm |
| CD7 | K036C2 | 176Yb | Biolegend* |
| CD16 | 3G8 | 209Bi | Fluidigm |
| Cell-ID Ir | n/a | Ir191 and Ir193 | Fluidigm |

\*metal conjugation performed in house

